# Rates of COVID-19-Associated Hospitalization in Immunocompromised Individuals in Omicron-era: A Population-Based Observational Study Using Surveillance Data in British Columbia, Canada

**DOI:** 10.1101/2022.08.22.22278955

**Authors:** Taraneh Bahremand, Jiayun Angela Yao, Christopher Mill, Jolanta Piszczek, Jennifer M. Grant, Kate Smolina

## Abstract

**Background:** People with immune dysfunction have a higher risk for severe COVID-19 outcomes. Omicron variant is associated with a lower rate of hospitalization but higher vaccine escape. This population-based study quantifies COVID-19 hospitalization rate in the Omicron-dominant era among vaccinated people with immune dysfunction, identified as clinically extremely vulnerable (CEV) population before COVID-19 treatment was widely offered.

**Methods:** All COVID-19 cases were reported to the British Columbia Centre for Disease Control (BCCDC) between January 7, 2022 and March 14, 2022. Case and population hospitalization rates were estimated across CEV status, age groups and vaccination status. Cumulative rates of hospitalizations for the study period were also compared between CEV and non-CEV individuals matched by sex, age group, region, and vaccination characteristics.

**Findings:** A total of 5,591 COVID-19 reported cases and 1,153 hospitalizations among CEV individuals were included. A third vaccine dose with mRNA vaccine offered additional protection against severe illness in CEV individuals. Vaccinated CEV population still had a significantly higher breakthrough hospitalization rate compared with non-CEV individuals.

**Interpretation:** CEV population remains a higher risk group and may benefit from additional booster doses and pharmacotherapy.

**Funding:** BC Centre for Disease Control and Provincial Health Services Authority

## Introduction

The Omicron variant of the severe acute respiratory syndrome coronavirus 2 (SARS-CoV-2) is clinically and epidemiologically different from previous variants. It is associated with higher transmissibility, increased immune escape, and less severe COVID-19 illness.^1^ During the Omicron wave in England, a lower proportion of COVID-19 hospitalizations were admitted primarily for COVID-19 compared to the Delta wave, where incidental diagnoses were less common.^2^ British Columbia (BC) is a province in Canada (population 5.2 million) where about 90% of individuals over 18 were vaccinated with at least 2 doses as of January 7^th^ 2022. The majority of the population received mRNA vaccines ((BNT162b2 [Pfizer-BioNTech] or mRNA-1273 [Moderna]). The first case of Omicron was identified in BC on 30 November 2021, and by end of December 2021, it became the dominant strain in BC.^3^

While vaccination is shown to be protective against severe COVID-19 ^4,5^ in the general population, people with compromised immune systems may produce less robust cell-mediated immunity^6^ and lower antibody levels, which may also decline faster over time.^7,8^ With previous variants, immunocompromised patients have fared worse clinically, but there is limited epidemiological information on COVID-19 severe outcomes during Omicron-era among immunocompromised populations. A recent study in the US found increased risk of breakthrough infection among immunocompromised individuals, independent of age, sex, and other comorbidities.^9^

A variety of therapeutics such as antivirals and monoclonal antibodies have been approved for treatment in patients with mild to moderate COVID-19. While these therapies have been shown to decrease the risk of hospitalization, they have not been specifically evaluated in immunocompromised individuals or against Omicron. The heavily mutated Omicron variant leads to decrease in neutralizing activity of many monoclonal antibodies and may be responsible for the decreased effectiveness,^10^ and antiviral treatments are costly, often available in limited supply and/or have the potential to cause adverse effects and drug-drug interactions. Therefore, it is important to accurately characterize the baseline epidemiological risk of severe disease of the intended recipients of these agents so that clinical and policy decisions can be made weighing the absolute benefit versus risk of pharmacotherapy, and to determine which patients should be prioritized for treatment.

As part of surveillance program at BC Centre for Disease Control (BCCDC), we aimed to assess the risk of hospitalization from COVID-19 among members of the population who meet the clinically extremely vulnerable (CEV) criteria. Depending on the pathophysiology and severity of conditions, CEV individuals were classified into three groups: severely immunocompromised (CEV Group 1), moderately immunocompromised (CEV Group 2), and complex conditions that were not immunocompromising but had a very high risk of hospitalization from COVID-19 (CEV Group 3).^11^ The CEV criteria are currently used in BC to prioritize patients for additional vaccine doses and early therapies for mild to moderate COVID-19 once therapies became available.

We report on the assessment on COVID-19 related breakthrough hospitalizations with consideration of age and vaccination status that was used to support decisions on prioritization of pharmacotherapy in BC.

## Materials and Methods

### Study population

Almost every resident of BC has public-funded healthcare service coverage and holds a unique Personal Health Number (PHN). Data on reported COVID-19 cases, hospitalizations, and vaccinations, as well as clinically extremely vulnerable (CEV) classification, were linked by PHN. Data on COVID-19 cases among adults aged 18 or over with a surveillance episode date (definition available in Data Source section) between January 7, 2022 and March 14, 2022 and hospitalizations with an admission date within two weeks of case surveillance episode date were included. Cases and hospitalizations were excluded if (1) age, CEV classification or vaccination status was unknown; or (2) cases were reported as re-infected according to the Public Health Reporting Data Warehouse reinfection algorithm; or (3) the surveillance health authority was recorded as “out of country”; or (4) residents or patients associated with an acute care facility, long term care facility or assisted living facility reportable outbreak (for hospitalization only); or (5) they had an admission date 3 or more days before the surveillance episode date, thus reflecting possible nosocomial infections. During this time period, the vast majority of COVID-19 infections were due to the Omicron variant.^12^

### Data Sources

#### COVID-19 vaccination

All COVID-19 vaccines administered in BC were recorded in the Provincial Immunization Registry (PIR),^13^ with information including the dose number, date of administration, trade name of the vaccine, as well as the recipients’ PHN, age, and regional health authority of residence.

#### COVID-19 cases and hospitalizations

COVID-19 cases, including lab-confirmed, lab-probable, and epi-linked cases, were reported to the BCCDC by all five regional health authorities in the province. Detailed case definitions can be found elsewhere.^14^ COVID-19 hospitalizations were reported by HAs as any case admitted to a hospital for at least an overnight stay, for reasons directly or indirectly related to their COVID-19 infection, and with no period of complete recovery between illness and admission. The data also include information on age, sex, regional health authority of residence, and case surveillance episode date, defined as symptom onset date; or earliest lab collection or result date if symptom onset date is not available; or public health case reported date if neither is available.

#### CEV classification

CEV individuals were defined using data-driven approach by National Advisory Committee on Immunization (NACI) having any of the underlying medical conditions listed in Table S1, and further classified into three groups of severely and moderately immunocompromised and those with complex conditions. Group 1 contains severely immunocompromised patients such as those with actively treated hematological malignancies, stem cell or solid organ transplant recipients; Group 2 includes moderately immunocompromised patients such as those with solid tumors, advanced HIV, or receiving certain immunosuppression therapies; Group 3 includes complex conditions associated with poor COVID-19 outcomes such as cystic fibrosis, developmental and intellectual disabilities and insulin-requiring diabetes. These individuals were identified primarily from their health care service records, and more detailed information about the definition and identification can be found elsewhere.^15^

### Vaccination status definition

The vaccination status of the study population at the time of infection was defined as follows: vaccinated, 3 doses: 14 days and more after administration of third dose; vaccinated, 2 doses: 14 days and more after administration of second dose AND less than 14 days after administration of third dose; vaccinated, 1 dose: 21 days or more after administration of first dose AND less than 14 days after administration of second dose; unvaccinated: no vaccination record OR less than 21 days after administration of first dose. A small fraction of the BC population received the Janssen (Johnson & Johnson) vaccine (0.001%). They were classified as “vaccinated, 2 doses” 21 days after first dose.

### Population and case hospitalization rates

A retrospective chart review of hospital records was done using a standard data extraction tool to better understand the distribution of reasons for admission for Omicron cases. It included BC residents who were over 18 years and had polymerase chain reaction (PCR) lab confirmed diagnosis of COVID-19. A stratified random sample of 600 hospital records from all five regional Health Authorities (HAs) reported lab results between 1^st^ December 2021 to 15^th^ January 2022 were reviewed. Hospital charts for those cases that were part of acute care outbreaks were included but represented about 5% of the patients. Results from this analysis were presented by the BC Provincial Health Officer at the media briefing.^16^

Chart review findings were then further stratified by CEV status used to produce estimates of percentage (%) of cases hospitalized primarily for COVID-19 (Table S2). These estimates were then subsequently applied to case hospitalization calculations to inform treatment prioritization groups. While case hospitalization metric in and of itself overestimates the true hospitalization risk because it only uses reported COVID-19 cases in the denominator and misses all other cases in the community who did not test, it still reflects critical patterns by age, vaccination, and CEV status. Furthermore, a positive test is the basis for determining treatment eligibility and represents a practical reference for that purpose.

### Matched analysis

To accompany the case hospitalization analyses, we also performed population-level matched analyses to confirm the observed patterns. CEV individuals were prioritized for vaccination in BC throughout the pandemic, and they generally received their vaccination earlier than the non-CEV population. They were also more likely to receive the Moderna vaccine, as per the national guidelines.^17^ To account for these potential confounding factors when comparing to a non-CEV population, a matched analysis was conducted. For each vaccinated CEV individual, five non-CEV individuals were randomly chosen who received the same number of doses, and had the same sex, 10-year age group, administration date of the last dose, trade name of the vaccine, number of vaccine doses, and health authority of residence. Cumulative rates of hospitalizations during the study period (first Omicron wave) were calculated and compared across the matched CEV and non-CEV groups.

### Statistical analysis

We performed a sensitivity analysis by excluding individuals who were treated with Paxlovid or Sotrovimab during the study period. All data processing, analysis, and visualization were done in R (Version 4.1.2). The confidence intervals in this study were calculated using exact method for Poisson rate, with the R package *epitools*.

## Results

Among the 311,818 adult CEV individuals in BC, 5,591 CEV individuals had a reported COVID-19 infection during the study period, of whom 1,153 were hospitalized. CEV population was notably older than non-CEV population (Table 1). The majority of CEV COVID-19 cases were in Group 2 (41%) or Group 3 (49%), reflecting the higher proportion of these individuals in the study population compared to CEV 1. Among CEV individuals, vaccine coverage was 88% for 2 doses and 51% for 3 doses at the beginning of the study period. Approximately 9% of CEV COVID-19 cases did not receive any type of COVID-19 vaccination as per their record.

**Table 1.**
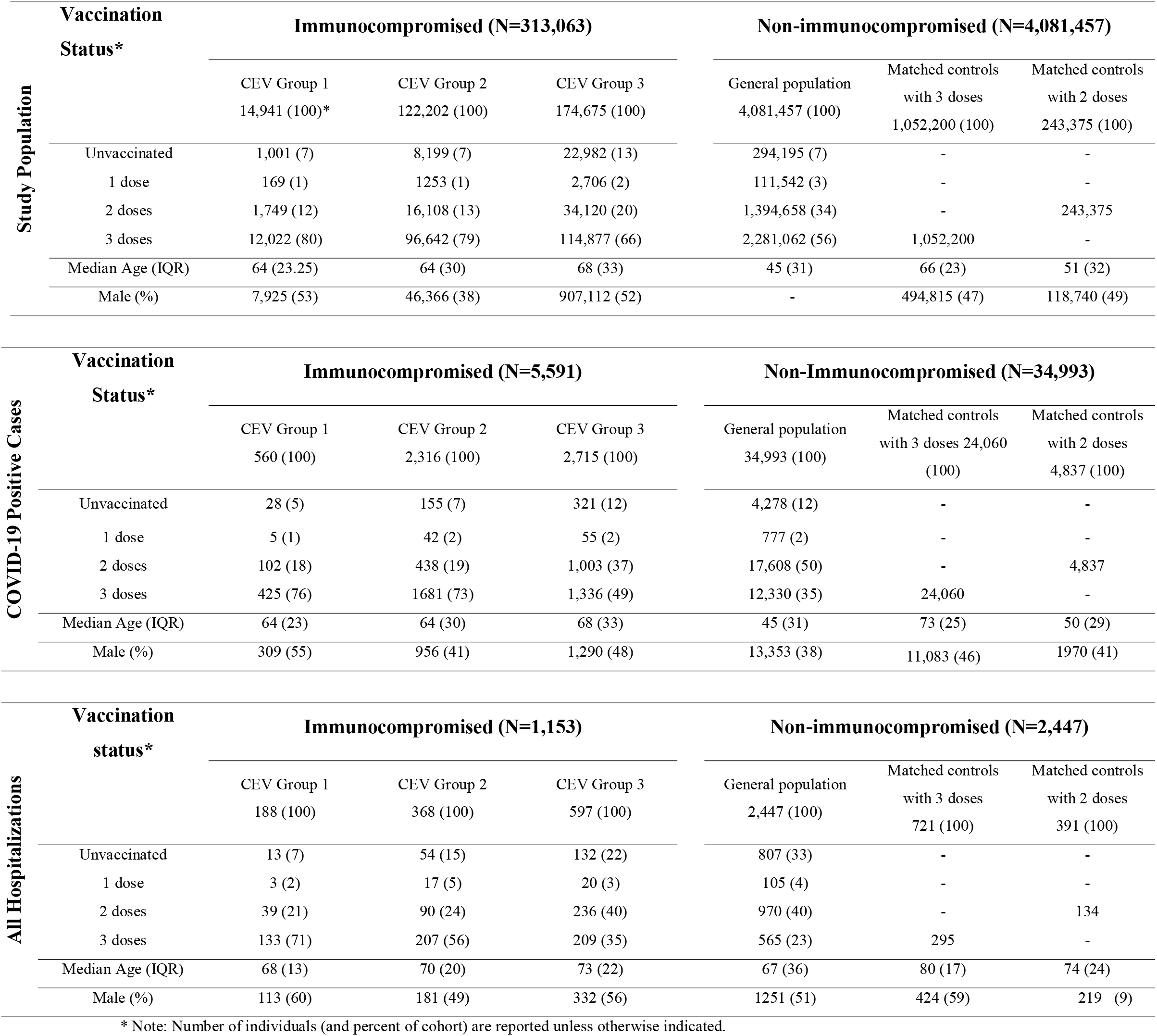
Baseline characteristics of the cohorts of study population, COVID-19 positive cases and all hospitalization according to the immunity and vaccine status.

## Discussion

In the context of Omicron predominance, we found that among both CEV and non-CEV populations, hospitalization rate was higher among older individuals and those with fewer vaccine doses. Vaccinated cases remained substantially less likely to be hospitalized compared to unvaccinated cases; this observation remained consistent in all groups regardless of the level of immunosuppression. Third dose offered additional protection against breakthrough hospitalization to everyone, regardless of the CEV status. However, people with immunocompromising conditions had a significantly higher rate of breakthrough COVID-19 hospitalizations compared with people without immunocompromising conditions.

Most clinical trials of COVID-19 vaccines excluded or underrepresented people with compromised immune systems, resulting in a limited amount of evidence on the impact of vaccination on this population. Here, we provide real-world evidence on hospitalization rates amid the period when Omicron was the dominant variant of concern. As Omicron is more prone to immune escape but associated with a lower likelihood of severe disease, these findings characterize COVID-19 epidemiology in the current pandemic context before treatment became widely available. Previous epidemiological studies in organ transplant recipients who received third doses of vaccination showed an improvement in the serum neutralizing activity against the Omicron variant. Our study provides evidence from real-world observations, in a more comprehensive immunocompromised population.

We also show that for each immune status, risk of hospitalization retains a strong independent age-gradient pattern, leading to much higher likelihood of severe disease among older individuals. This finding is consistent with previous studies in BC and other jurisdictions,^18,19^ which have shown age as one of the major predictors of severe COVID-19.

Among CEV population, CEV group 1 saw significantly higher hospitalization rate compared with CEV group 2 and group 3. This is expected, because group 1 includes the most immunocompromised people with higher baseline risk of hospitalization rate and higher rate of vaccine failure.^20,21^ The CEV groups 2 and 3 represent a wide range of conditions treated with immunosuppressive medications. The CEV 3 group is a broader umbrella in which some members may be minimally or not immune suppressed, but remain at risk for severe disease. While we did not observe notable differences between the two groups, it is important to emphasize that they include a heterogeneous mix of conditions and there is variation in the risk of severe outcomes between different conditions.

Various therapeutic agents shown to reduce disease progression and prevent hospitalization in clinical trials are now available for early treatment of COVID-19. Selecting the appropriate candidates for therapy is challenging as these trials were conducted in unvaccinated individuals before the emergence of Omicron and had low external generalizability to the current pandemic situation, when most of the adult population is vaccinated and the dominant variant is Omicron. These treatments are also not harmless, with the key drug, Paxlovid, having multiple drug interactions and potentially substantial side effects. This study shows key relationships between age, vaccine status, and immune status to help prioritize patients at highest risk of severe disease in whom treatments may have the most benefit. In British Columbia, CEV populations have been prioritized for treatment on the basis of these findings.

Our study has a number of strengths. It is one of the first population-based observational studies reporting breakthrough hospitalization estimates specific to immunocompromised individuals while adjusting for potentially incidental findings of COVID-19, a key feature that distinguishes Omicron-era epidemiology from prior variants. Our analysis was conducted using data in a large universal health care system with a diverse and stable population, benefitting from comprehensive capture of COVID-19 vaccinations, testing, and disease outcomes. Unlike vaccine effectiveness studies that focus on relative metrics (such as odds ratios or hazard ratios), we provide absolute population and case rate of hospitalization, which allows for a concrete quantification of risk that is necessary for clinical, operational, and logistical aspects of treatment eligibility decision making.

There are several limitations. Due to changes in immunocompromising conditions or medications, there is some risk of misclassification to and from the study groups as some people may not have met the criteria for CEV definitions used in this study throughout the whole study period, but we expect this number to be small. Testing rates were higher among CEV individuals compared with non-CEV individuals, reflecting, at least in part, BC’s “test-to-treat” strategy to navigate appropriate use of limited antiviral treatment for high-risk COVID-19 positive cases. This means that otherwise younger and healthy adults would not have been eligible for publicly funded PCR testing, while those at higher risk, including CEV individuals, were prioritized. As such, results likely overestimate case hospitalization rate in the Not CEV group; the relative difference in case hospitalization rates between CEV and non-CEV populations is likely even greater. Case and population level hospitalization rate calculations do not account for time since last vaccine dose, but we address this limitation in the matched analysis. We assumed equal risk of exposure to SARS-CoV-2 in the matched CEV and non-CEV individuals, but this assumption may not always hold. The non-CEV population who were vaccinated at the same time as the CEV population might have some special characteristics for prioritization, such as being health care workers, which may present different risk of exposure to the virus than the general population. Although this study was done with the entire population of a well-defined geographic region, it is possible that we missed some individuals who were not in the province for the full study period or misclassified vaccination status for those who received vaccination outside of the province.

The higher risk of breakthrough hospitalizations among CEV individuals who received 3 doses compared with non-CEV individuals suggests that the CEV population remains a higher risk group and may benefit from additional booster doses and pharmacotherapy. Our findings also highlight the need to continuously monitor health outcomes among immunocompromised individuals. Further studies should assess the risk-benefit of pharmacotherapy following infection among the CEV and non-CEV populations.

## Data Availability

Data is used for public health surveillance and cannot be shared by the authors. Data access request should be made to the data stewards (BC MOH and PHSA) as per existing processes. R codes for data processing, analysis and visualization can be shared upon request.

## Contribution

KS conceived the study idea. KS, CM, and JAY designed the study. CM and JAY performed the data analyses. TB performed the literature review and wrote the first draft of the manuscript. JP and JMG provided clinical consultations. All authors contributed to revising or critically reviewing the article. All authors gave final approval of the version to be published. All authors have agreed on the journal to which the article has been submitted and agree to be accountable for all aspects of the work.

## Ethics approval

This work was performed under BCCDC’s mandate to perform population health surveillance and falls under the Behavioural Research Ethics Board at the University of British Columbia (approval # H20-02097).

## Acknowledgements

We would like to acknowledge Lillian Ding, Ognjenka Djurdjev, Joanne Shum, Alexandra Flatt, and Dr. Maureen O’Donnell for providing CEV information and supporting this work. We also acknowledge Dr. Hind Sbihi and BC Public Health Laboratories for providing testing data. Our gratitude also goes to the individuals involved in health authority chart review and analysis to produce estimates of the proportion of hospitalizations primarily due to COVID-19: Sara Miles, Monica Durigon, Elisabeth Hanson, Dr. Jae Ford, Mei Chong, Dr. Felicity Clemens, Marsha Taylor, Dr. Caren Rose, Vancouver Coastal Health Public Health Surveillance Unit, Dr. Naveed Janjua and Dr. Jat Sandhu.

---

**Figure 1.**
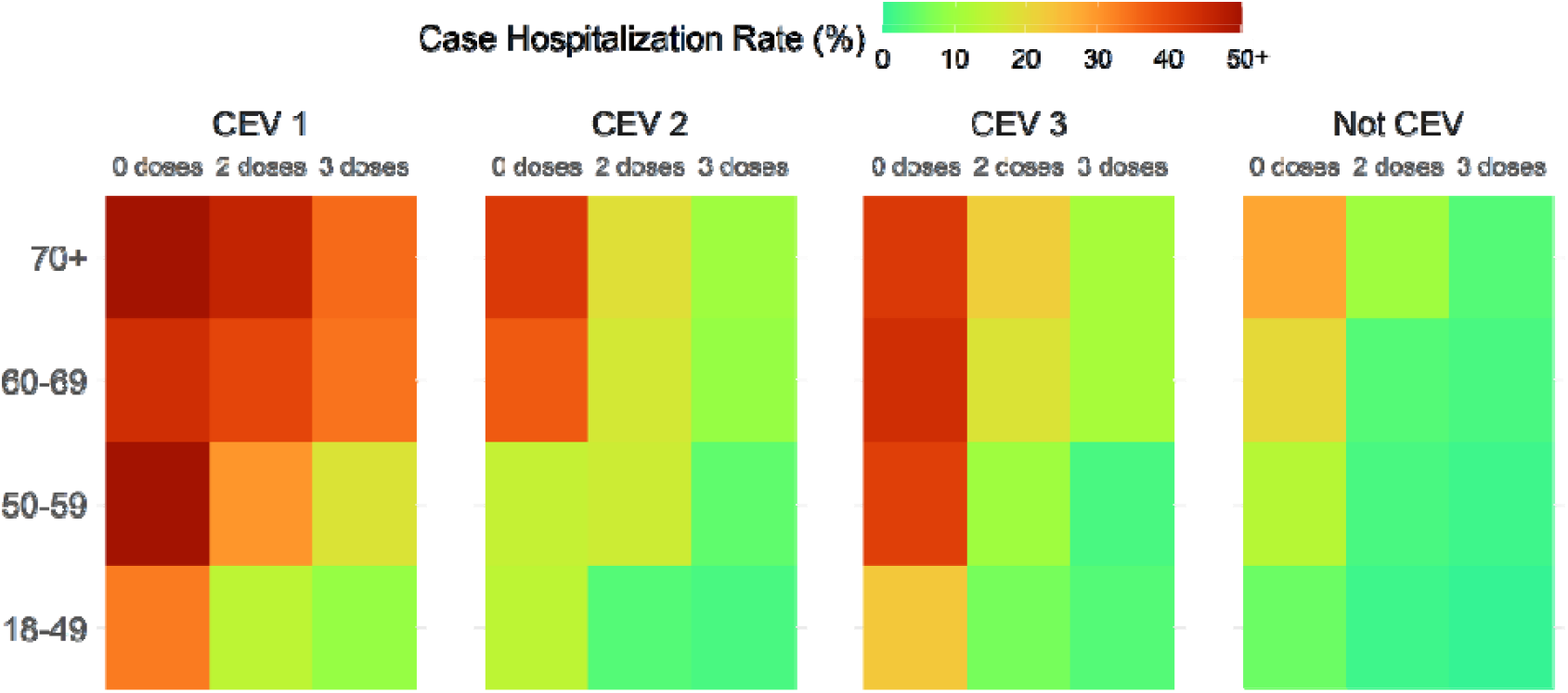
Heat map of the hospitalization rate (%) among reported COVID-19 cases by age group and their CEV and vaccination status. ***Figure 1*** shows the heat map corresponding to adjusted *case* hospitalization rate, shown as percentage, among vaccinated and unvaccinated groups, stratified by age group and CEV group. Each rectangle represents a group of interest, and its colour visualizes the estimated percent of cases who were hospitalized primarily due to COVID-19. *It is critical to note that this represents hospitalizations among reported cases only and thus greatly overestimates the true hospitalization rate among all infections that actually occurred*. The main objective of the figure is to illustrate the relative patterns by CEV group, age and vaccination status – not to accurately depict risk of severe outcomes. It reflects higher rates of hospitalization among older or/and unvaccinated people in all subgroups. It also illustrates the protective effect conferred by 3^rd^ dose of vaccine, both in CEV and non-CEV populations. Because reported case hospitalization rates are so much higher among the CEV population, the colour scheme is heavily skewed towards that group, making it harder to see the age-
 and dose- gradient in the non-CEV population.

**Figure 2.**
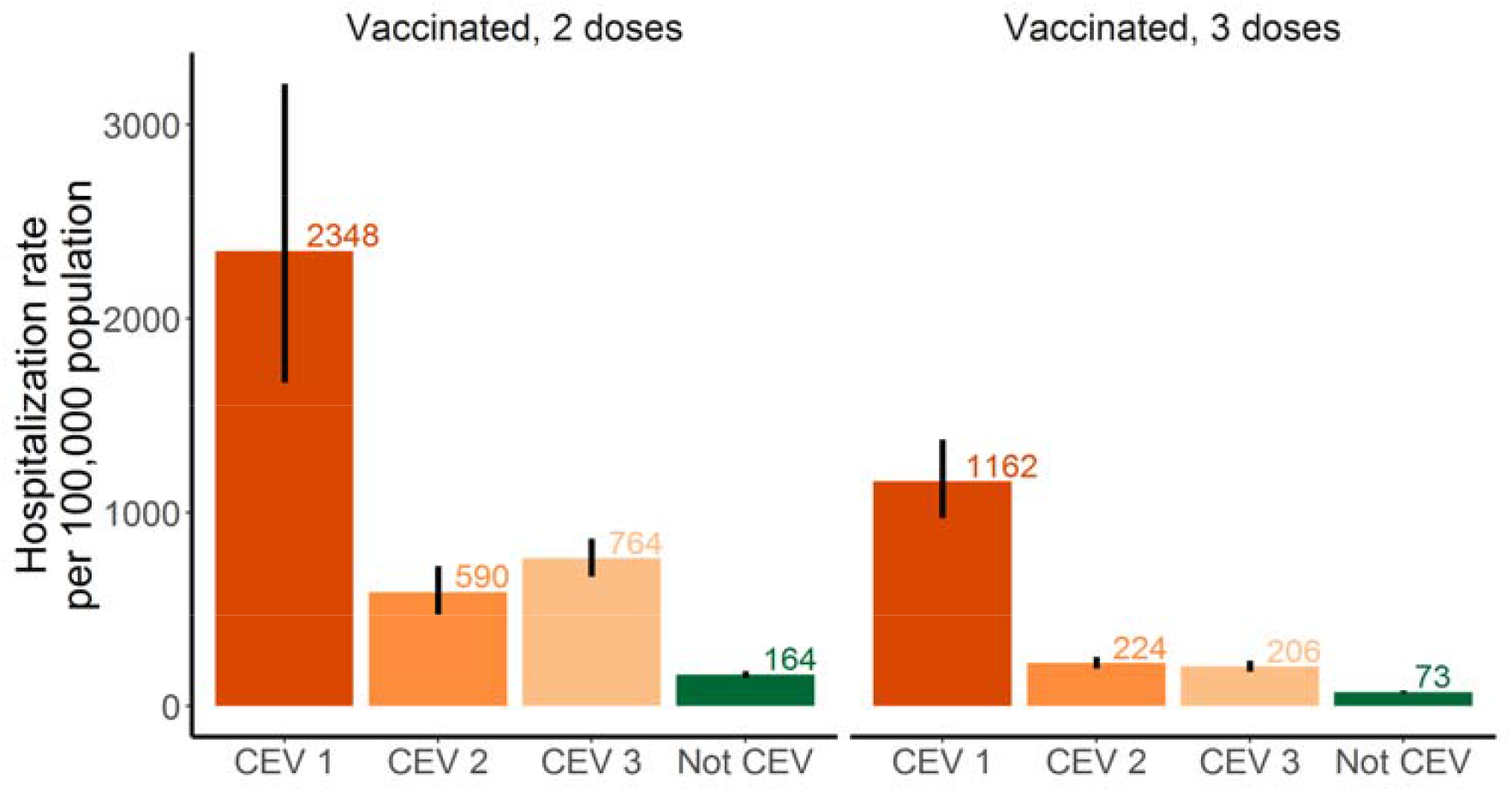
Breakthrough hospitalization rate per 100,000 among vaccinated immunocompromised (CEV) cases compared to their non-immunocompromised (Not CEV) matched individuals. ***Figure 2*** illustrates population-level rate of COVID-19 hospitalizations among vaccinated CEV individuals compared to matched vaccinated non-CEV individuals over the first two months of Omicron predominance in BC. In each CEV group and among non-CEV individuals, hospitalization rates were higher among 2 dose recipients compared to 3 dose recipients, similar to the case hospitalization findings illustrated in Figure 1. Among CEV individuals, hospitalization rates were significantly higher in Group 1, who are severely immunocompromised, compared with Groups 2 and 3, regardless of the number of vaccine doses. Exclusion of individuals who received Paxlovid or Sotrovimab (n=821) revealed minimal impact on the results displayed in Figure 1 and Figure 2 (data not shown).

