## Supplemental tables for "Rates of COVID-19-Associated Hospitalization in Immunocompromised Individuals in Omicron-era: A Population-Based Observational Study Using Surveillance Data in British Columbia, Canada"

**Table S1. CEV conditions and its classification into three severely and moderately immunocompromised and mild immunosuppression/complex conditions**

| **CEV Group 1** | |
|---|---|
| **Definition** | **Notes** |
| Solid Organ Transplant (SOT) recipients: Solid organ transplant recipients of kidney, liver, lung, heart, pancreas or islet cell, bowel or combination transplant. | |
| Those being actively treated for hematological malignancy: Have received or are receiving active treatment (chemotherapy, targeted therapies including CAR-T, immunotherapy) for malignant hematologic conditions (e.g., leukemia, lymphoma, or myeloma). | Have received treatment for haematological malignancy in the last year These medications may not come up on PharmaNet as they are administered in hospital facilities such as BC Cancer |
| Those who have had a bone marrow or stem cell transplant: Have had bone marrow or stem cell transplant or are still taking immunosuppressant medications related to transplant | Pregnant and have a serious heart disease, congenital or acquired, that requires observation by a cardiac specialist throughout pregnancy<br>Pregnant and have a serious heart disease, congenital or acquired, that requires observation by a cardiac specialist throughout pregnancy |
| Those who have taken anti-CD20 agents or B-cell depleting agents which cause suppression of their immune response:<br>i. Have received treatment with any anti-CD20 agents (i.e., rituximab, ocrelizumab, ofatumumab, obinutuzumab, ibritumomab, tositumomab) or<br>ii. B-cell depleting agents (i.e., epratuzumab, MEDI-551, belimumab, BR3-Fc, AMG-623, Atacicept, anti-BR3, alemtuzamab). | Pregnant and have a serious heart disease, congenital or acquired, that requires observation by a cardiac specialist throughout pregnancy |
| Those with severe primary immuno-deficiencies: Have combined immune deficiencies affecting T-cells, immune dysregulation (particularly familial hemophagocytic lymphohistiocytosis) or those with type 1 interferon defects (caused by a genetic primary immunodeficiency disorder or secondary to anti-interferon autoantibodies). | There are less than 100 individuals in this category in BC |
| **CEV Group 2** | |
| **Definition** | **Notes** |
| Those additional patients who have received treatment for cancer including solid tumors: | Systemic cancer therapy received in the last 6 months<br>Radiation received in the last 3 months |

| | |
|---|---|
| i. Have received or are receiving systemic therapy (including chemotherapy, molecular therapy, immunotherapy, targeted therapies including CAR-T, monoclonal antibodies, hormonal therapy for cancer other than the hematological malignancies in CEV 1;<br>i ii. Have received or are receiving radiation therapy for cancer. | |
| Those who have taken significantly immunosuppressing drugs:<br>i. Anti-CD20 agents: rituximab, ocrelizumab, ofatumumab, obinutuzumab, ibritumomab, tositumomab;<br>ii. B-cell depleting agents: epratuzumab, MEDI-551, belimumab, BR3-Fc, AMG-623, Atacicept, anti-BR3, alemtuzamab;<br><br>iii. Biologics: abatacept, adalimumab, anakinra, benralizumab, brodalumab, canakinumab, certolizumab, dupilumab, etanercept, golimumab, guselkumab, infliximab, interferon products (alpha, beta, and pegylated forms), ixekizumab, mepolizumab, natalizumab, omalizumab, resilizumab, risankizumab, sarilumab, secukinumab, tildrakizumab, tocilizumab, ustekinumab, or vedolizumab; iv. Oral immune-suppressing drugs: azathioprine, baricitinib, cyclophosphamide, cyclosporine, leflunomide, dimethyl fumerate, everolimus, fingolimod, mycophenolate, siponimod, sirolimus, tacrolimus, tofacitinib, upadacitinib, methotrexate, or teriflunomide; iv. Oral steroids on an ongoing basis: dexamethasone, hydrocortisone, methylprednisolone, or prednisone;<br>iv. vi. Immune-suppressing infusions/injections: cladribine, cyclophosphamide, glatiramer, methotrexate | Anti-CD20 agents taken in the last 2 years<br>B-cell depleting agents taken in the last 2 years<br><u>Anti-CD20 and B-cell depleting agents may not be on PharmaNet</u><br>Biologics taken in the last 3 months<br>Oral immunosuppressing drugs taken in the last month<br>Oral steroids equivalent to 20mg/d of prednisone equivalent (adult dose) taken on an ongoing basis in the last month<br><u>Infusions/injections in vi. taken in the last 3 months</u> |
| Those with advanced untreated HIV infection or those with acquired immuno-deficiency syndrome (AIDS) defined as AIDS defining illness or CD4 count ≤ 200/mm3 or CD4 fraction ≤ 15% | Untreated HIV or treated HIV with CD4 count ≤ 200/mm3 qualifies the patient for treatment but referral to an HIV Specialist is recommended due to complexity of patients and drug-drug interactions. However, treatment with nirmatrelvir/ritonavir should not be withheld or delayed |
| People with moderate primary immunodeficiencies: Have a moderate to severe primary immunodeficiency which has been diagnosed by an adult or pediatric immunologist and requires ongoing immunoglobulin replacement therapy (IVIg or SCIG) or the primary | IVIg and SCIg treatment will not be visible on PharmaNet<br><u>There are <1000 such patients in BC</u> |

| | |
|---|---|
| immunodeficiency has a confirmed genetic cause (e.g., DiGeorge syndrome, Wiskott-Aldrich syndrome). | |
| Those with on dialysis and those with severe kidney/renal disease:<br>i. Dialysis (hemodialysis or peritoneal dialysis);<br>ii. Stage 5 chronic kidney disease (eGFR <15ml/min);<br>iii. Glomerulonephritis and receiving steroid treatment | Patients with renal disease are not eligible to receive nirmatrelvir/ritonavir as it is contraindicated in severe renal disease. Sotrovimab should be used in patients with renal disease. Criteria for sotrovimab are: |
| **CEV Group 3** | |
| **Definition** | **Notes** |
| Patients with severe respiratory disorders:<br>i. Cystic fibrosis,<br>ii. Severe COPD: hospitalized because of COPD<br>iii. Severe asthma: hospitalized because of asthma<br>iv. Are taking biologics for asthma, severe lung disease and at least one of the following: long-term home oxygen; assessment for a lung transplant; severe pulmonary arterial hypertension; severe pulmonary fibrosis/interstitial lung disease. | Hospitalized for COPD in the last year Hospitalized for asthma in the last year Taking biologics in the last 3 months |
| Rare blood disorders:<br>Homozygous sickle cell disease, highest risk thalassemia (Received an attestation letter. The full definition is: a diagnosis of thalassemia and two of the following: transfusion dependent; receiving iron chelation therapy; pre-transfusion hemoglobin levels <70 in last 2-3 years; have iron overload; have had a splenectomy or have other significant health conditions; are over 50, Atypical Hemolytic Uremic Syndrome or Paroxysmal Nocturnal Hemoglobinuria) | There are <200 people in BC with these disorders. The full definition is for completeness. Patients will have received communication from Bonnie Henry if their rare blood disorder qualifies. |
| Rare metabolic disorders: certain metabolically unstable inborn errors of metabolism: urea cycle defects; methylmalonic aciduria; propionic aciduria; glutaric aciduria; maple syrup urine disease. | There are only 87 patients in this category |
| Had a splenectomy: Anatomical or functional asplenia | |
| Diabetes treated with insulin | This is the largest CEV category and comprises of both Type 1 and Type 2 diabetes |
| Hematological and other cancers not captured in CEV group 1 or 2 | This is a very broad category: patients would have received communication about their CEV status.<br>It is important to distinguish whether the patient with cancer fits into category 1, 2 or 3 as vaccine status is not a consideration for treatment in category 1 and 2, whereas only patients who are unvaccinated or partially vaccinated would qualify for treatment in CEV 3.<br>Examples of cancers here are chronic hematological malignancies under surveillance (e.g., chronic |

| | |
|---|---|
| | lymphocytic leukemia) or active solid tumours not on treatment but undergoing surveillance. |
| Significant developmental disabilities: Down Syndrome, or Cerebral Palsy, or Intellectual Developmental Disability (IDD), or receiving supports from:<br>• Community Supports for Independent Living (CSIL) or<br>• Community Living British Columbia (CLBC): currently receiving supports or assessed and eligible for CLBC supports or<br>• Nursing Support Services program for youth aged 16 and above<br>• People aged 12+ whose condition is described but are not using support services can receive priority through consultation with their health-care provider (attestation form) | |
| Pregnant and have a serious heart disease, congenital or acquired, that requires observation by a cardiac specialist throughout pregnancy | Reproductive Infectious Diseases specialist on call at BCWH can be consulted for assistance with this group as needed as no therapy is specifically approved in pregnancy. |
| Pregnant and have a serious heart disease, congenital or acquired, that requires observation by a cardiac specialist throughout pregnancy | |

### Table S2. Estimated % hospitalizations that are primarily due to COVID-19 based on results of health authority chart review

| Age group | CEV Group 1 | | CEV Groups 2 & 3 | | Not CEV | |
|---|---|---|---|---|---|---|
| | 0 doses | 1+ doses | 0 doses | 1+ doses | 0 doses | 1+ doses |
| 18-49 | 90% | 50% | 80% | 40% | 50% | 20% |
| 50+ | 100% | 90% | 90% | 60% | 70% | 40% |
